# Ancestrally and Temporally Diverse Analysis of Penetrance of Clinical Variants in 72,434 Individuals

**DOI:** 10.1101/2021.03.11.21253430

**Authors:** Iain S. Forrest, Kumardeep Chaudhary, Ha My T. Vy, Shantanu Bafna, Daniel M. Jordan, Ghislain Rocheleau, Ruth J.F. Loos, Judy H. Cho, Ron Do

## Abstract

A major goal of genomic medicine is to quantify the disease risk of genetic variants. Here, we report the penetrance of 37,772 clinically relevant variants (including those reported in ClinVar^1^ and of loss-of-function consequence) for 197 diseases in an analysis of exome sequence data for 72,434 individuals over five ancestries and six decades of ages from two large-scale population-based biobanks (Bio*Me* Biobank and UK Biobank). With a high-quality set of 5,359 clinically impactful variants, we evaluate disease prevalence in carriers and non-carriers to interrogate major determinants and implications of penetrance. First, we associate biomarker levels with penetrance of variants in known disease-predisposition genes and illustrate their clear biological link to disease. We then systematically uncover large numbers of ClinVar pathogenic variants that confer low risk of disease, even among those reviewed by experts, while delineating stark differences in variant penetrance by molecular consequence. Furthermore, we ascertain numerous variants present in non-European ancestries and reveal how increasing carrier age modifies penetrance estimates. Lastly, we examine substantial heterogeneity of penetrance among variants in known disease-predisposition genes for conditions such as familial hypercholesterolemia and breast cancer. These data indicate that existing categorical systems for variant classification do not adequately capture disease risk and warrant consideration of a more quantitative system based on population-based penetrance to evaluate clinical impact.

---

The advent of high-throughput sequencing has led to an exponential increase in genetic tests for health-related purposes^2,3^. The American College of Medical Genetics & Genomics (ACMG) recommends clinical action if a pathogenic genetic variant is found in one of 59 genes (hereafter ACMG59)^4^. This “genetics-first” approach is feasible if variant pathogenicity is ascertained with high confidence and veracity. A large database of human variation and phenotypes, ClinVar^1^, classifies variant pathogenicity in a scheme that informs clinical interpretation of genetic test results (e.g., “pathogenic”, “uncertain significance”, “benign”)^5^. However, most ClinVar variants are of uncertain clinical significance, and misclassification of 12% of pathogenic and 90% of conflicting variants has inflated their pathogenicity^6^. Variants implicated in diseases from breast cancer to cardiomyopathy have been scrutinized for overestimated claims of pathogenicity^7^ and many variants classified as pathogenic were recently downgraded to lower ClinVar classes^8^. Any unreliability of pathogenicity, already a categorical rather than quantitative metric of disease risk, further diminishes its clinical value. There is therefore a critical need to provide accurate information for a variant’s effect on disease to guide patient care.

Penetrance, the probability of a disease phenotype given a particular genotype^9^, is unknown for the vast majority of variants in ClinVar and genetic databases. Well-known exceptions include ∼60% of carriers of certain *BRCA1* and *BRCA2* variants develop breast cancer by 70 years of age^10^ and 73% of individuals heterozygous for *LDLR* variants present with hypercholesterolemia^11^. Findings of highly penetrant variants immediately guide treatment, such as statins, and screening, such as mammograms^12,13^. Penetrance thus gives meaningful and actionable information for variants by quantifying a carrier’s disease risk.

Until recently, penetrance has been primarily derived from family-based or clinical cohort studies^14–17^. These typically focus on a small number of genes and inherently maximize penetrance estimates by recruiting patients with disease or family history of disease, and are therefore susceptible to ascertainment bias^18^. Small sample size and genetic or environmental modifiers further limit their ability to reliably appraise penetrance^19^. In contrast, the recent introduction of the UK Biobank (UKB) and other population-based biobanks has created a trove of genetic and phenotypic data for large numbers of unrelated individuals^20,21^. Exome sequences coupled to electronic health records (EHRs) provide an unprecedented opportunity for using a population-based method to measure penetrance on a large scale with less ascertainment bias than traditional studies^22^. In addition, multi-ethnic biobanks such as Mount Sinai’s Bio*Me* Biobank (Bio*Me*)^23^ enable the analysis of penetrance across diverse ancestries. Recent studies have begun to use a population-based approach to explore penetrance of genes in breast cancer^24^ and familial hypercholesterolemia (FaH)^25^, yet the population-based penetrance of most variants, genes, and diseases remains uncharacterized.

Here, we perform a comprehensive analysis of variant penetrance observed in two large-scale EHR-linked population-based biobanks (Bio*Me* and UKB). We measured the penetrance of 37,772 variants, including a high-quality set of 5,359 clinically impactful variants, for 197 diseases of dominant inheritance using exome sequences from 72,434 individuals. Penetrance was analyzed in all individuals and separately in five ancestries across six decades of ages. We investigated determinants of penetrance, including ClinVar pathogenicity, variant molecular consequence, and carrier ancestry and age. The clinical manifestation of variants was scrutinized by examining metabolite levels, biological measurements, and physician notes for carriers, and clinically significant variants and genes were highlighted. This highly scalable, population-based approach of generating penetrance estimates for thousands of clinically relevant variants expands our knowledge of the genetic influences on disease and has the potential to advance genomic medicine.

## RESULTS

### Analysis of penetrance of clinical variants

A graphical abstract is provided (**Supplementary Figure 1**). Whole-exome sequencing, quality control, and filtering were performed on 80,773 samples from two population-based EHR-linked biobanks (**Methods**) to generate a final dataset with 72,434 individuals (**Table 1**). Demographics, self-reported ancestry, biological measurements, and International Classification of Disease 10 (ICD-10) diagnosis codes were available for individuals in the final dataset. We defined case-control status for over 400 non-recessive diseases in ClinVar sourced from the Systematized Nomenclature of Medicine Clinical Terms (SNOMED CT)^26^ using ICD-10 codes^27^, of which 197 had at least one case in the final dataset (**Supplementary Table 1**). We analyzed 37,772 variants that were either reported in ClinVar for at least one of the aforementioned diseases or were of predicted loss-of-function (LoF) consequence (**Supplementary Table 2**). A stringent set of 5,359 clinically impactful (impactful) variants—defined by pathogenic/likely pathogenic classification in ClinVar or LoF annotation (splice acceptor/donor, stop gained/lost, frameshift, start lost) with Variant Effect Predictor^28^ in a gene mediating disease via LoF mechanism, and in a gene with non-recessive inheritance—was used for downstream analyses unless otherwise stated (**Supplementary Figure 2**). We identified carriers with at least one allele (hereafter referred to as carriers) for each variant and computed the proportion of carriers affected with disease to determine penetrance. Variant penetrance was estimated in all carriers and separately in carriers of five diverse ancestries and six age ranges. The proportion of non-carriers affected with disease was also quantified for each variant to ascertain disease prevalence among the population in the absence of a scorable variant.

**Table 1.** Overview of baseline demographic and clinical traits for 72,434 individuals from two population-based biobanks.

| Trait | BioMe<br>(n=29,039) | UKB<br>(n=43,395) |
| --- | --- | --- |
| Male, n (%) | 11,684 (40%) | 19,330 (45%) |
| Age, mean (SD) | 59 (16) | 57 (8) |
| European ancestry, n (%) | 9,376 (32%) | 40,447 (93%) |
| African ancestry, n (%) | 7,190 (25%) | 916 (2%) |
| Hispanic ancestry, n (%) | 8,528 (29%) | -- |
| Asian ancestry, n (%) | 1,349 (5%) | 1,088 (3%) |
| Mixed ancestry, n (%) | 2,028 (7%) | 344 (0.8%) |
| BMI in kg/m2, mean (SD) | 28 (7) | 28 (5) |
| SBP in mmHg, mean (SD) | 127 (20) | 140 (19) |
| DBP in mmHg, mean (SD) | 73 (12) | 82 (11) |
| LDL-C in mg/dL, mean (SD) | 98 (36) | 63 (16) |
| Total cholesterol in mg/dL, mean (SD) | 177 (45) | 102 (21) |
| HbA1c in %, mean (SD) | 6 (2) | 6 (3) |
| Glucose in mg/dL, mean (SD) | 106 (47) | 93 (20) |
| HTN, n (%) | 12,962 (45%) | 11,094 (26%) |
| T2D, n (%) | 7,044 (24%) | 2,820 (6%) |
| FBC, n (%) | 1,488 (5%) | 1,581 (4%) |
| AMD, n (%) | 1,041 (4%) | 352 (0.8%) |

Overview of baseline demographic and clinical traits for 72,434 individuals from two population-based biobanks: BioMe Biobank (BioMe) and UK Biobank (UKB). n, number; SD, standard deviation; SBP, systolic blood pressure; DBP, diastolic blood pressure; LDL-C, low-density lipoprotein cholesterol; HbA1c, hemoglobin A1c; HTN, hypertension; T2D, type 2 diabetes; FBC, familial breast cancer; AMD, age-related macular degeneration; --, no Hispanic ancestry individuals are in UKB.

### Validation of phenotyping and penetrance approach

We performed a series of analyses to validate our population-based approach of phenotyping and computing penetrance. First, we selected nine diseases from a variety of systems—age-related macular degeneration (AMD), arrhythmogenic right ventricular cardiomyopathy (ARVC), FaH, familial breast cancer (FBC), type 2 diabetes (T2D), etc.—and identified cases using both ICD-10 diagnosis codes and previously published clinical algorithms^29–37^. We then evaluated and compared the penetrance of 208 ClinVar pathogenic variants corresponding to the diseases in Bio*Me* using both phenotype approaches (**Supplementary Table 3**). Similar penetrance values were observed for all diseases. Three diseases (Brugada syndrome, hepatocellular carcinoma, and pulmonary arterial hypertension) had equivalent mean variant penetrance with both approaches; mean variant penetrance of the other six diseases varied by 8% or less. A tabulated list of these results is provided (**Supplementary Table 4**).

Second, we further assessed the accuracy of our phenotyping approach by manually reviewing physician notes in the problem list (PL) for cases and controls of six representative diseases: Alzheimer’s disease (AD), AMD, FaH, FBC, stroke, and T2D. We randomly sampled 50 ICD-10-based cases and controls for each disease and examined their PL for symptoms or findings indicative of a diagnosis while blinded to ICD-10 disease status (**Supplementary Table 5**). We observed high concordance in case-control classification between ICD-10- and PL-based phenotyping, ranging from 100% (for AD) to 93% (AMD, FaH). Case concordance ranged from 88% (AMD, FaH)– 100% (AD, FBC, T2D), while control concordance ranged from 94% (T2D)–100% (AD, stroke).

### Biomarker levels linked to variant penetrance

We investigated metabolite and clinical measurements of carriers of impactful variants with different penetrance in genes with clear biological links to three diseases: FaH^12^, maturity-onset diabetes of the young (MODY)^38^, and obesity^39,40^ (**Figure 1**). Carriers of penetrant variants for FaH in *LDLR* had greater low-density lipoprotein cholesterol (LDL-C) and total cholesterol levels than carriers of incompletely and nonpenetrant variants (**Figure 1a**). Using linear regression, we observed robust association between variant penetrance and LDL-C (effect size [β]=0.53 mg/dL per 1% increase in penetrance, standard error [SE]=0.12; *P*=0.004) and total cholesterol levels (β=0.98 mg/dL, SE=0.18; *P*=0.002) adjusting for clinical covariates and statin use. Analogously, carriers of penetrant variants for MODY in hepatocyte nuclear factor-1α (*HNF1A*) had elevated glucose and hemoglobin A1c (HbA1c) levels compared to carriers of incompletely penetrant variants (**Figure 1b**). Variant penetrance was significantly associated with glucose (β=0.76 mg/dL, SE=0.19; *P*=0.008) and HbA1c levels (β=0.083% [2.4 mg/dL], SE=0.020 [0.57 mg/dL]; *P*=0.01) adjusted for clinical covariates and diabetes medication use. Lastly, carriers of penetrant variants for obesity in uncoupling protein 3 (*UCP3*) had higher body mass index (BMI) than carriers of incompletely and nonpenetrant variants (**Figure 1c**). Variant penetrance was strongly associated with BMI (β=0.24 kg/m^2^, SE=0.044; *P*=6×10^-4^) and weight (β=1.3 lbs [0.59 kg], SE=0.26 [0.12 kg]; *P*=8×10^-4^) adjusting for clinical covariates. Together, these analyses using published phenotype algorithms, physician notes, and biological measurements support the validity of our population-based penetrance method and illustrate the clinical presentation of impactful variants.

**Figure 1.**
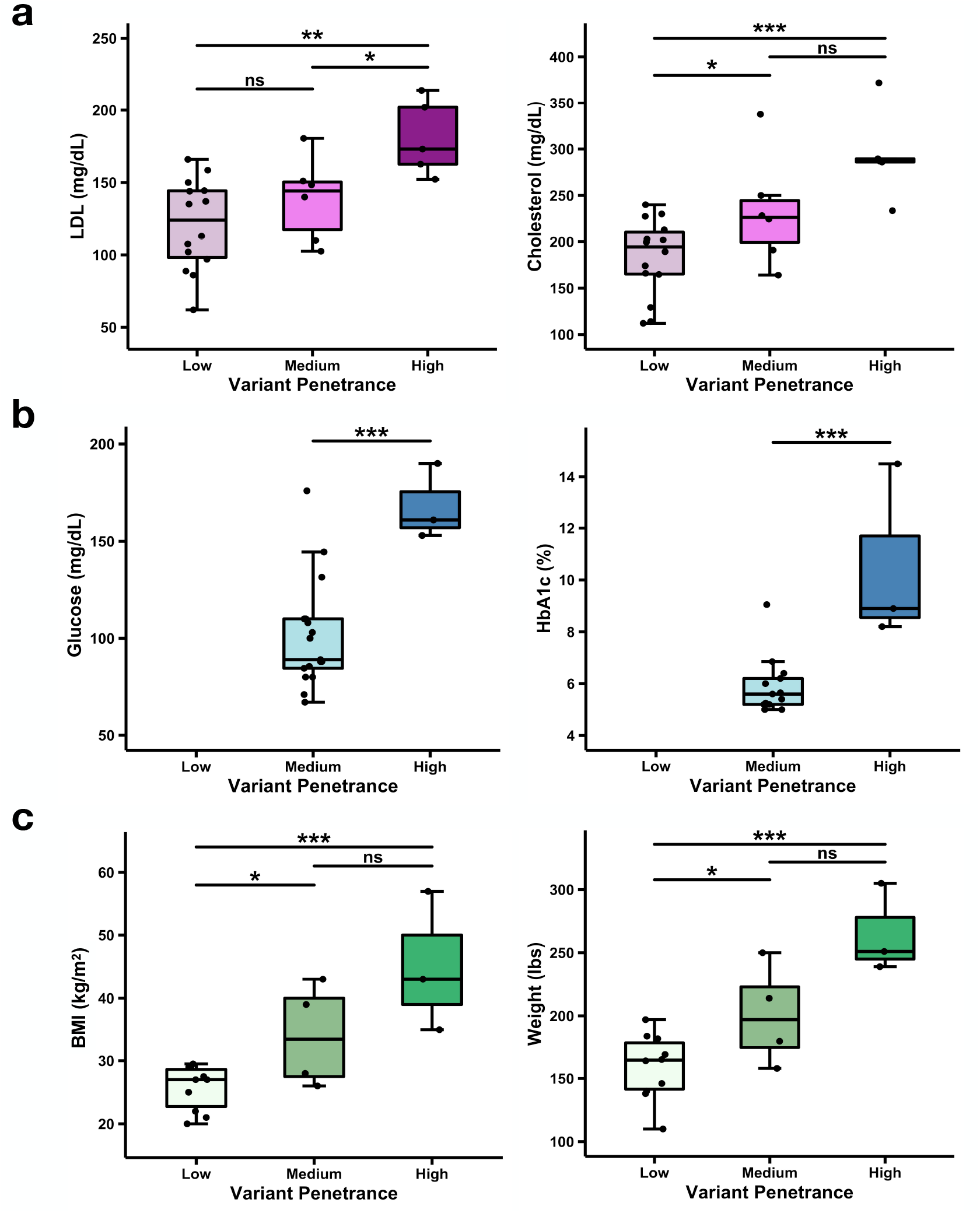
Biomarker levels in carriers of clinically impactful variants with varying penetrance for three diseases in the Bio*Me* Biobank. Metabolite and clinical measurements for carriers of clinically impactful variants with varying penetrance for three diseases in the Bio*Me* Biobank. Clinically impactful variants are reported as pathogenic/likely pathogenic in ClinVar or are loss-of-function in a gene that mediates disease via loss-of-function mechanism, and in a gene with non-recessive inheritance. Mean of measurements is compared between penetrance strata with two-tailed t-test: *, *P*<0.05; **, *P*<0.01; ***, *P*<0.001. **a**, Variants in low-density lipoprotein receptor (*LDLR*) are stratified by penetrance for familial hypercholesterolemia (Low [0%], Medium [greater than 0 and less than 100%], and High [100%]) and shown as box plots with measurements of low-density lipoprotein cholesterol (LDL) and total cholesterol (cholesterol) levels in carriers. **b**, Variants in hepatocyte nuclear factor 1-α (*HNF1A*) are stratified by penetrance for maturity-onset diabetes of the young (Low [0%], Medium [greater than 0 and less than or equal to 50%], and High [greater than 0 and less than 100%]) and depicted as box plots with levels of glucose and hemoglobin A1c (HbA1c) in carriers. No impactful variants in *HNF1A* were of Low penetrance. **c**, Variants in uncoupling protein 3 (*UCP3*) are stratified by penetrance for obesity (Low [0%], Medium [greater than 0 and less than 100%], and High [100%]) and displayed as box plots with body mass index (BMI) and weight of carriers.

### Distribution of observed variant penetrance and disease risk

We examined the distribution of disease risk conferred by 5,359 impactful variants and 157 corresponding diseases. Risk difference (RD) is the difference between the prevalence of disease in carriers and non-carriers, and represents the excess risk of disease attributed to the variant of interest. A summary of mean variant RD for all 157 diseases is provided (**Supplementary Table 6**). We observed 565 (11%) variants with RD exceeding 0.05 for 55 (35%) diseases (**Figure 2**). In contrast, we detected a very large number of weakly penetrant and nonpenetrant variants that confer little to no disease risk (4,794 [89%] variants with RD≤0.05), which can be attributed in part to ClinVar pathogenic variants from traditional studies that maximize penetrance estimates and have biased ascertainment^18^. Recent studies have demonstrated overestimation of the disease rates of these variants^6–8^, indicating a need for population-based approaches to more unbiasedly gauge penetrance.

**Figure 2.**
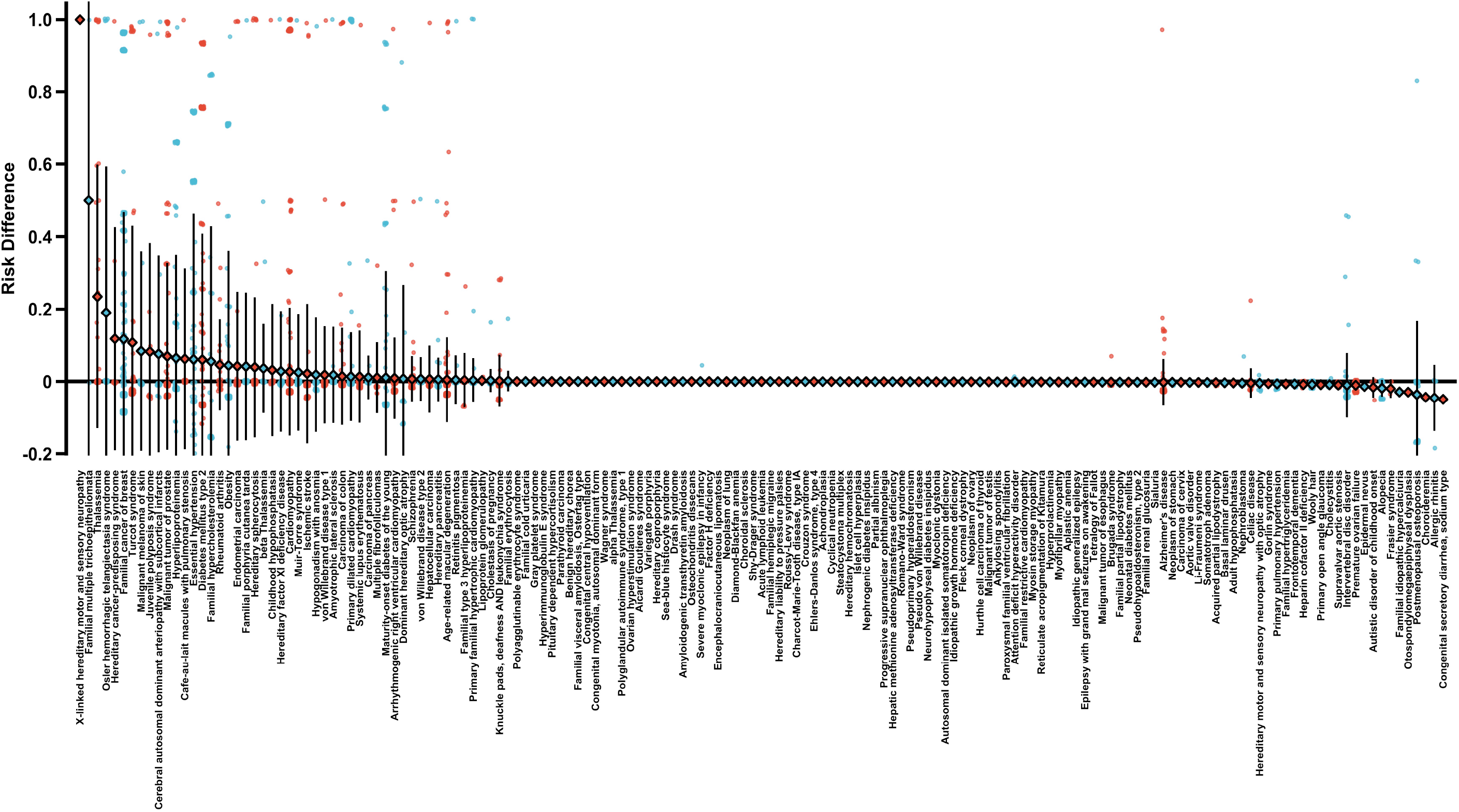
Distribution of observed disease risk for 5,359 clinically impactful variants in 72,434 individuals. Distribution of observed disease risk for 5,359 clinically impactful variants in 72,434 individuals. Risk difference is the difference in disease prevalence between carriers and non-carriers of a variant of interest. Clinically impactful variants are reported as pathogenic/likely pathogenic in ClinVar or are loss-of-function in a gene that mediates disease via loss-of-function mechanism, and in a gene with non-recessive inheritance. Diseases are sorted by descending mean variant risk difference (diamonds), with all variant risk difference estimates plotted (points) along with the standard deviation (error bars) per disease. A complete tabulated list of results is provided in **Supplementary Table 6**.

Next, we investigated the prevalence of disease in carriers (i.e., penetrance) and non-carriers of impactful variants according to ClinVar pathogenicity (pathogenic, uncertain, conflicting, benign) (**Figure 3a**), ClinVar review status (expert reviewed, multiple submitters, single submitter, no assertion criteria) (**Figure 3b**), and molecular consequence (**Figure 3c**). We hypothesized that variants reported as pathogenic in ClinVar, reviewed by experts, or with LoF consequence have greater *a priori* evidence of disease risk and are expected to have higher penetrance than variants reported as benign, without criteria assertion, or of synonymous consequence. In agreement with these expectations, we observed that mean variant penetrance was highest among pathogenic (6.9% vs. 0.86% next highest mean for uncertain class; *P*=5×10^-270^), expert reviewed (18% vs. 6.4% next highest mean for multiple submitters; *P*=9×10^-17^), and frameshift variants (10% vs. 4.1% for missense; *P*=2×10^-13^). The mean penetrance of ClinVar variants with conflicting (0.75%), uncertain (0.86%), and benign clinical significance (0.085%) was similar, warranting caution when interpreting variants that do not fully meet the ACMG’s strict pathogenic criteria^41^. We also note that the mean penetrance of ClinVar pathogenic variants was only 6.9% and only 18% for those with the highest review status (expert reviewed), indicating that these broad classifications do not adequately capture all variants with large effect on disease as measured by penetrance.

**Figure 3.**
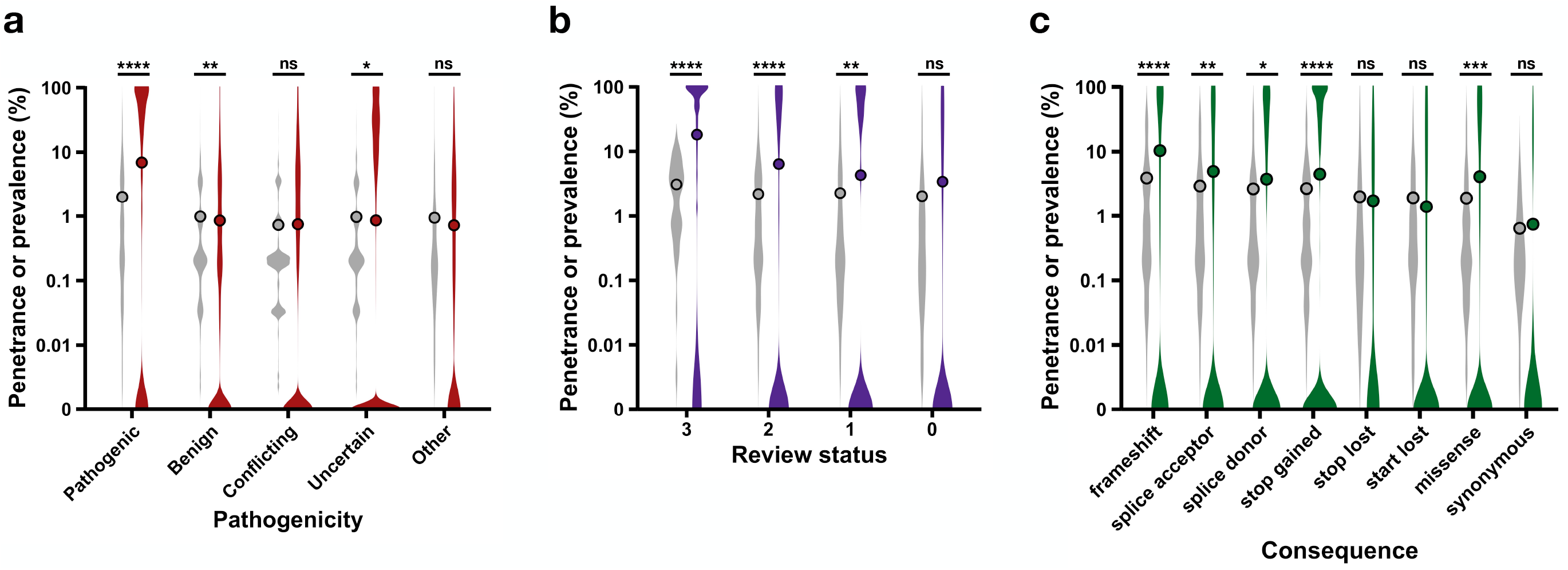
Penetrance of 34,301 variants stratified by ClinVar pathogenicity, ClinVar review status, and molecular consequence. Penetrance of 34,301 variants in non-recessive genes stratified by ClinVar pathogenicity, ClinVar review status, and molecular consequence. Penetrance distributions are shown as violin plots in red, purple, and green color on a base-10 logarithmic scale with the mean penetrance overlaid as points, alongside disease prevalence in non-carriers shown as violin plots in grey color with the mean disease prevalence superimposed as points. Pathogenic/likely pathogenic variants are grouped as pathogenic and benign/likely benign variants are grouped as benign. *, two-tailed t-test *P*<0.05; **, *P*<0.01; ***, *P*<0.001; ****, *P*<0.0001. **a**, Penetrance is stratified by classification of variant pathogenicity in ClinVar. Pathogenic variants on average have the highest penetrance (6.9% vs. 0.86% next highest mean penetrance in uncertain class; *P*=5×10^-270^). Only pathogenic variants confer a significantly increased risk of disease on average to carriers compared to baseline disease risk in non-carriers (risk difference [RD]=0.046; *P*=1×10^-22^). Other pathogenicity, variants with a ClinVar pathogenicity classification other than pathogenic, benign, conflicting, or uncertain. **b**, Penetrance is stratified by variant review status in ClinVar as reviewed by experts (review status=3), multiple submitters (review status=2), single submitter (review status=1), or no assertion criteria (review status=0). Variants reviewed by experts (review status=3) have the highest penetrance on average (18% vs. 6.3% next highest mean penetrance in review status=2 group; *P*=9×10^-17^). Pathogenic variants of review status=3, =2, and =1 confer a significantly increased risk of disease on average, with RD=0.15 (*P*=1×10^-13^), RD=0.042 (*P*=3×10^-10^), and RD=0.020 (*P*=4×10^-3^), respectively. **c**, Penetrance is stratified by molecular consequence annotated with Variant Effect Predictor (VEP). Frameshift variants on average have the highest penetrance (10% vs. 4.1% for missense; *P*=2×10^-13^). There was a significantly elevated risk of disease on average with frameshift (RD=0.064; *P*=3×10^-19^), splice acceptor (RD=0.020; *P*=5×10^-3^), splice donor (RD=0.011; *P*=4×10^-2^), stop gained (RD=0.018; *P*=3×10^-7^), and missense variants (RD=0.022; *P*=4×10^-4^).

We conducted several sensitivity analyses to account for differing sample sizes of penetrance estimates. Penetrance distributions according to ClinVar pathogenicity, review status, and molecular consequence remained similar when stratified by increasing thresholds of sample sizes (**Supplementary Figure 3**). Singletons (very rare variants appearing only once) comprised a large portion of the impactful variants (3,507/5,359 [65%]) and statistically are completely penetrant or nonpenetrant. To test the validity of singleton penetrance, we evaluated the proportion of penetrant singletons by ClinVar pathogenicity, review status, and molecular consequence (**Supplementary Figure 4**). If singleton penetrance were accurate, the proportion of penetrant singletons among pathogenic, expert reviewed, or LoF singletons would be expected to exceed that of benign, non-expert reviewed, or non-LoF singletons, respectively. We observed that the proportion of penetrant singletons was in fact greatest among pathogenic (13% vs. 2.6% in next highest class of conflicting; *P*=2×10^-19^), expert reviewed (41% vs. 12% in next highest group of multiple submitters; *P*=4×10^-11^), and frameshift singletons (16% vs. 7.8% for missense; *P*=1×10^-3^). We therefore retained penetrance estimates of smaller sample sizes in our analyses.

### Ancestral and temporal dimensionality of penetrance data

Most penetrance studies to date have focused on one ancestry, typically European^42,43^, with few notable exceptions^11,23,44^. Yet it is well-known that allele frequency (AF) and disease risk of variants can vary substantially in different populations^45,46^. Thus, we computed penetrance among multiple self-reported ancestries, including European, African, Hispanic, and Asian ancestries. To demonstrate this diversity, we identified numerous ancestry-specific impactful variants. Out of 2,209 impactful variants in Bio*Me*, we observed 75 (3.4%), 43 (1.9%), 17 (0.77%), and 71 (3.2%) variants exclusively present in African, Hispanic, Asian, and European ancestry, respectively. Out of 3,408 impactful variants in UKB, we observed 8 (0.23%), 11 (0.32%), and 235 (6.9%) variants specific to African, Asian, and European ancestry, respectively. Furthermore, we identified ancestry-specific variants that were highly penetrant, such as an Asian ancestry-specific pathogenic frameshift variant in *HBB* (NC_000011.10:c.5226994_5226995insC) associated with a greatly increased risk of thalassemia (RD=0.99; *P*=9×10^-6^) and a European ancestry-specific pathogenic frameshift variant in *PALB2* (NC_000016.9:c.23647358_23647359del) associated with a significantly elevated risk of breast cancer (RD=0.92; *P*=0.007).

We also delineated variant penetrance based on different age thresholds of carriers ranging from at least 20 years (≥20) to at least 70 years (≥70). Age of disease onset is pertinent for estimating penetrance: congenital or early onset diseases will have manifested in older carriers of penetrant variants, whereas later onset diseases may not have presented yet in younger carriers. To address temporality, we characterized the observed change in variant penetrance with increasing carrier age for 157 diseases corresponding to the impactful variants, stratified by age of disease onset (**Figure 4**; **Supplementary Figure 5**). We first annotated and grouped the diseases by age of onset: Earlier (congenital, childhood, or adolescent), Later (adulthood), or Any (**Supplementary Table 7**). For each disease group, we then calculated the change in variant penetrance (ΔPenetrance) between the lowest carrier age threshold and increasing age thresholds (e.g., variant penetrance for ≥20 versus ≥30, ≥20 versus ≥40, etc.). As expected, the mean ΔPenetrance increased in the Later disease group with higher carrier age thresholds in Bio*Me* (**Figure 4a**). The largest mean ΔPenetrance was +1.8% in the Later disease group when comparing ≥20 and ≥70, while mean ΔPenetrance remained ∼0% in the Earlier and Any disease groups. In UKB, no significant differences in mean ΔPenetrance were observed in the Later disease group (**Figure 4b**) likely due to its older age range of carriers (40-69 years) whereby Later diseases would have manifested. Thus, penetrance estimates of diseases with early or any age of onset remained stable over time, while penetrance estimates of diseases with later age of onset predictably increased with higher carrier ages.

**Figure 4.**
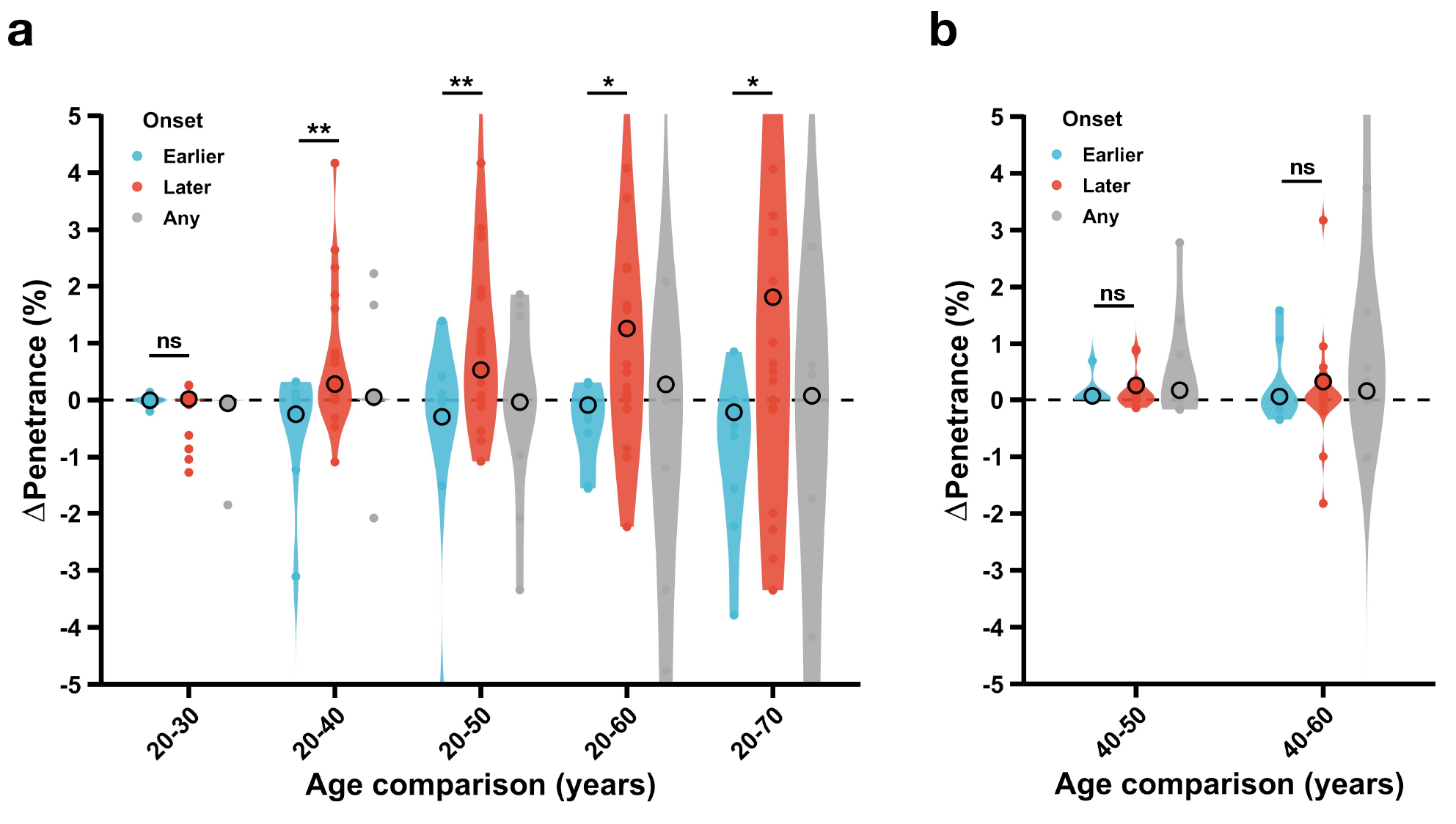
Association between age of disease onset and age-dependent change in penetrance for 157 diseases. Association between age of disease onset and age-dependent change in penetrance for 157 diseases. Diseases correspond to the 5,359 clinically impactful variants and are grouped according to their age of onset: Earlier, Later, or Any. Change in penetrance is displayed as a violin plot for each age of onset group when comparing two carrier age thresholds with the mean change in penetrance superimposed as a point. ΔPenetrance (%), change in variant penetrance represented as a percent (+ values indicate greater penetrance estimate with the older age threshold and - values indicate greater penetrance estimate with the younger age threshold); age comparison, two carrier age thresholds for which penetrance is compared (e.g., 20-70 compares penetrance with carriers ≥20 years old and penetrance with carriers ≥70 years old); onset, disease groups according to age of onset; *, two-tailed t-test *P*<0.05; **, *P*<0.01; ***, *P*<0.001; ****, *P*<0.0001. A complete plot including diseases with ΔPenetrance >5% or ΔPenetrance <-5% is provided (**Supplementary Figure 5**). **a**, The Bio*Me* Biobank has carriers of age 20-90 years old and ΔPenetrance is assessed over five carrier age thresholds: Later diseases have greater ΔPenetrance on average than Earlier diseases for 20-40 (*P*=0.006), 20-50 (*P*=0.002), 20-60 (*P*=0.02), and 20-70 (*P*=0.02) age comparisons. **b**, UK Biobank has carriers of age 40-69 years old and ΔPenetrance is evaluated over two carrier age thresholds: there is no difference in ΔPenetrance between Later and Earlier diseases for either 40-50 (*P*=0.2) or 40-60 (*P*=0.2) age comparisons.

### Clinical utility of variant penetrance data

A genotype-first approach in genomic medicine must identify penetrant variation in clinically significant genes to inform screening, management, and treatment. The ACMG recommends reporting secondary findings of pathogenic variants in the ACMG59, but acknowledges that insufficient data on penetrance requires ongoing study and revision^49^. *BRCA1* and *BRCA2* are both in the ACMG59, and a recent study estimated the population-based penetrance of pathogenic variants in these genes for FBC using data from the UKB^50^. However, penetrance was estimated at the gene level, and non-ClinVar variants were omitted as were variants in other genes that have been shown to confer significant risk of breast cancer, such as *PALB2*, *CHEK2*, *ATM*, *PTEN*, and others^24^. Thus, we investigated the penetrance of impactful variants in 10 known or suspected breast cancer-predisposition genes (*BRCA1*, *BRCA2*, *PALB2*, *CHEK2*, *ATM*, *PTEN*, *CDH1*, *BARD1*, *BRIP1*, and *RAD51D*) to illustrate the clinical value of a population-based approach of determining penetrance at the variant level (**Figure 5a**). The highest disease risk on average was conferred by impactful variants in *BRCA1* (mean variant penetrance=38%, mean RD=0.32; P=2×10^-6^), *BRCA2* (mean variant penetrance=38%, mean RD=0.32; *P*=1×10^-10^), and *PALB2* (mean variant penetrance=26%, mean RD=0.20; *P*=0.009).

**Figure 5.**
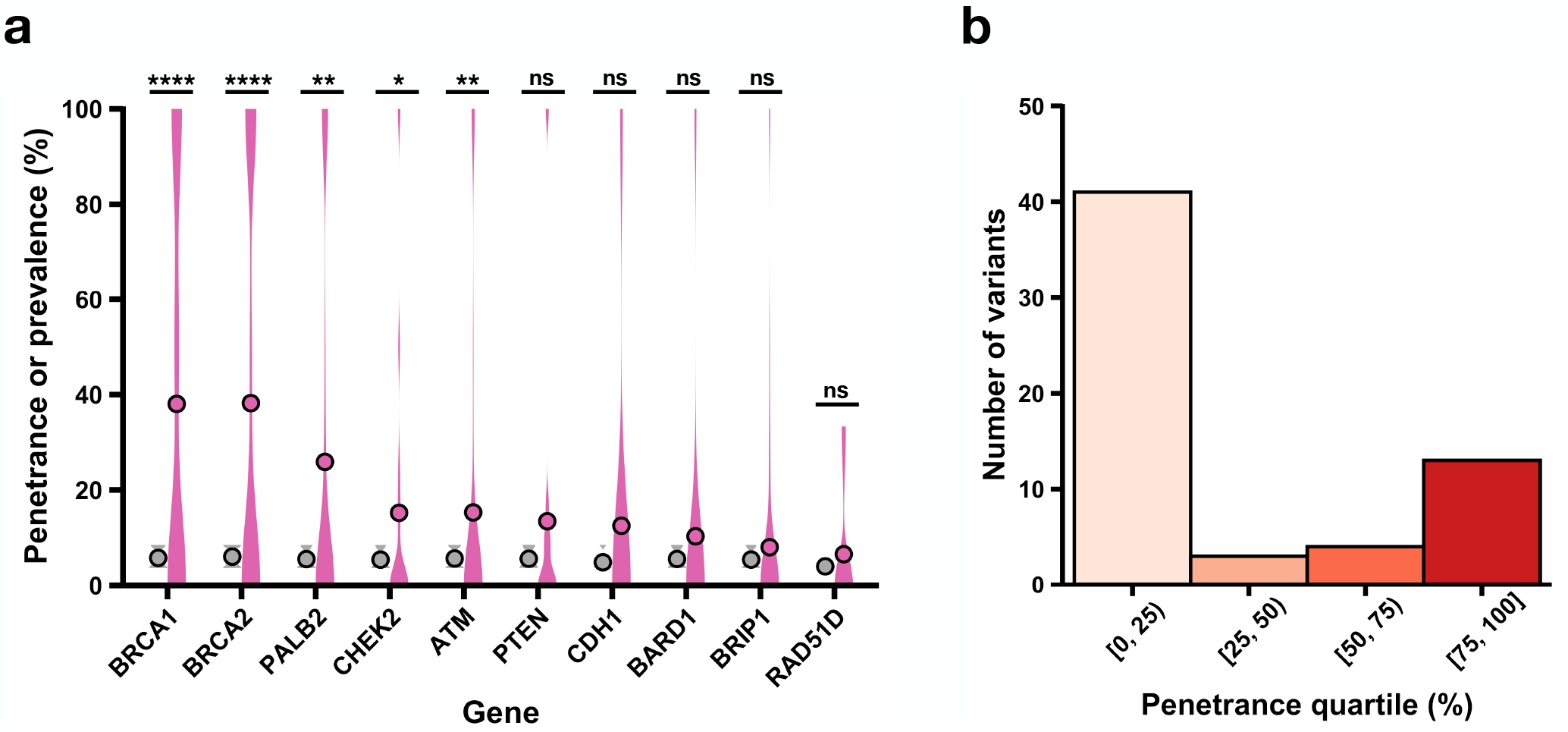
Disease risk associated with clinically impactful variants in disease-predisposition genes for breast cancer and hypercholesterolemia. Disease risk associated with clinically impactful variants in known breast cancer- and hypercholesterolemia-predisposition genes. Clinically impactful (impactful) variants are reported as pathogenic/likely pathogenic in ClinVar or are loss-of-function in a gene that mediates disease via loss-of-function mechanism, and in a gene with non-recessive inheritance. **a**, Penetrance of impactful variants in 10 known or suspected breast cancer-predisposition genes is displayed as pink violin plots with the mean penetrance overlaid as a point, alongside disease prevalence in non-carriers shown as grey violin plots with the mean prevalence superimposed as a point. Violin plots are sorted by genes with descending mean penetrance, with the highest mean variant penetrance of 38% in *BRCA1* (n=48 variants, mean risk difference [RD]=0.32; *P*=2×10^-6^) and 38% in *BRCA2* (n=92 variants, mean RD=0.32; *P*=1×10^-10^). *, two-tailed t-test *P*<0.05; **, *P*<0.01; ***, *P*<0.001; ****, *P*<0.0001. **b**, Heterogeneity in the penetrance of 59 impactful low-density lipoprotein receptor (*LDLR*) variants is shown. The number of variants is plotted as a histogram across four quartiles of penetrance. [0, 25), penetrance less than 25%; [25, 50), penetrance greater than or equal to 25% and less than 50%; [50, 75), penetrance greater than or equal to 50% and less than 75%; [75, 100], penetrance greater than or equal to 75%.

These data clarify the disease risk of previously reported variants in ClinVar and novel variants not yet reported in ClinVar or the literature. We highlighted examples of variants that are significantly associated with risk of seven diseases, including FBC, to demonstrate this dual utility (**Supplementary Table 8**). Many impactful variants in *BRCA2* were strongly associated with FBC, such as a known pathogenic frameshift variant (NC_000013.11:c.32340301del; penetrance=47%, RD=0.39; *P*=9×10^-6^) and a previously unreported frameshift variant (NC_000013.11:c.32340630_32340631del; penetrance=100%, RD=0.96; *P*=4×10^-13^). Evidence of high penetrance strengthens a candidate variant’s case for clinical significance, while assertions of pathogenicity are refined by quantitative disease risk estimates.

Notably, we observed substantial heterogeneity in the penetrance of variants even within the same disease-predisposition gene. Impactful variants in *LDLR*, for instance, exhibited a wide range of penetrance for FaH (**Figure 5b**). Similarly, there was a large distribution of penetrance associated with impactful variants in *BRCA1* (variant penetrance standard deviation [SD]=45%, RD SD=0.44), *BRCA2* (variant penetrance SD=46%, RD SD=0.46), and *PALB2* (variant penetrance SD=42%, RD SD=0.42) for FBC (**Figure 6a**). Instead of coarsely categorizing variants as simply pathogenic versus non-pathogenic and collapsing all variants in a gene, these data reveal granularity and nuance in the penetrance of individual variants.

## DISCUSSION

A major goal of genomic medicine is to tailor clinical care of patients to their unique genetic composition, especially penetrant variants. Scalability and accuracy are crucial for determining penetrance and employing its information in a clinical setting. Conventional penetrance estimates from family-based or clinical cohort studies typically focus on one variant or gene at a time^14–17^ (limited scalability), and have small sample size, ascertainment bias, inconsistent carrier screening, and genetic/environmental confounders^18,19^ (limited accuracy). A proliferation of population biobanks, such as UKB and Bio*Me*, has made available thousands of genetic and phenotypic records^20,21,23^. This raises the possibility of using a population-based method to measure penetrance whereby large numbers of unrelated carriers are assessed^22^. Here, we performed a comprehensive assessment of variant penetrance using 72,434 exomes from two large-scale population-based biobanks, with the dataset freely accessible and provided in **Supplementary Tables 9** and **10**.

Reliable penetrance estimates depend on reliable phenotyping of carriers. We performed robust validation analyses to support our phenotyping and penetrance strategy. First, we computed penetrance for nine diseases using both diagnosis codes and previously published clinical algorithms, finding high concordance in all diseases tested. Second, we manually curated physician notes for six diseases to verify ICD-10-based diagnoses. Third, we extracted laboratory and clinical test results (e.g., levels of lipids, glucose, HbA1c, and BMI) for carriers and observed that penetrance was strongly associated with relevant quantitative traits, adjusted for medications and clinical covariates. Hence, by validating our phenotyping and penetrance method, and evaluating penetrance for thousands of variants in a high-throughput manner, we ensure both accuracy and scalability. As penetrance studies grow, similar validation analyses should be implemented to certify fidelity of penetrance estimates.

While several efforts have begun to probe the upward bias of penetrance estimates in traditional studies, there has not yet been a systematic investigation of the pervasiveness of overestimated penetrance. In the present study, we examined the distribution of disease risk associated with a high-quality set of 5,359 impactful variants and 157 diseases. Persistently lower disease risk was observed for many diseases, with a few exceptions of highly penetrant variants such as in *BRCA1* and *BRCA2*. This can be interpreted in light of a few important considerations: 1) small sample size and stochasticity for rarer variants may contribute to variable penetrance estimates; however, a number of analyses supported the accuracy of our penetrance measurements with respect to sample size, including for singletons (**Supplementary Figure 3**, **Supplementary Figure 4**); 2) ascertainment of cases in conventional studies has been shown to inflate the disease risk of variants^18,19^ and recent population-based studies with lower penetrance estimates for FaH and developmental disorders bolster this explanation^11,22^; 3) reported pathogenicity does not equate with penetrance, as we found by interrogating whether existing qualitative classification systems in ClinVar capture the quantitative disease risk of variants. ClinVar pathogenic variants were more penetrant than variants of other ClinVar classes yet still weakly penetrant overall, in line with previous studies^6–8^ and consistent with ClinVar’s definition of pathogenic that includes “low penetrance” variants (https://www.ncbi.nlm.nih.gov/clinvar/docs/clinsig/). We then examined a major driver of variation in penetrance among ClinVar pathogenic variants: when stratified by ClinVar review status (evidence for clinical significance of a variant), pathogenic variants that were not expert reviewed had lower penetrance.

In light of these findings, it is incumbent to discuss whether variants reported as pathogenic but empirically shown to have low penetrance should be classified differently, or whether categorical systems of disease risk (pathogenic versus non-pathogenic) should be complemented with a quantitative system based on penetrance^7^. Recent commentary by Khera and Hegele proposed a new paradigm for classifying FaH based on two parameters: carrier status for a pathogenic *LDLR* variant and severity of hypercholesterolemia^51^. This advanced schema for disease classification, and others like it, would greatly benefit from knowledge of penetrance for pathogenic *LDLR* variants to better stratify disease risk and personalize medical care.

We also emphasize the importance of including diverse ancestries and age ranges when characterizing variant penetrance. Populations differ by AF and disease factors^45,46^, yet most genetic studies have focused on Europeans^52,53^. Here, we capture ancestry-specific penetrance in detail, identifying over a hundred variants in non-European ancestries. While past studies have reported age-dependent penetrance in age-related diseases such as FBC^17,54^, amyotrophic lateral sclerosis^55,56^, and obesity^57^, these typically estimate gene-based penetrance whereby all variation in a gene is aggregated. In contrast, we evaluate age-dependent penetrance at the variant level. As expected, early onset diseases showed stable penetrance over time while later onset diseases had increasing penetrance with older carriers. For example, the observed penetrance of an expert reviewed ClinVar pathogenic frameshift variant in *BRCA2* (NC_000013.11:c.32340301del) increased from 38% in European ancestry carriers ≥40 years of age to 56% in those ≥60 years of age.

There were several study limitations. First, ICD-10 diagnosis codes from the EHR were used to define case-control status^26,27^ (**Supplementary Table 1**). While commonly used in EHR-linked biobank studies^61,62^, there may be some misclassification^63–66^. Though several validation analyses reinforced the soundness of our phenotyping, these were completed for a subset of the diseases. Second, we cannot exclude the possibility of potential bias in our datasets. Bio*Me* is predominantly composed of individuals recruited from the Mount Sinai Health System and may have a higher burden of diseases and therefore penetrance estimates. In contrast, the preponderance of healthy volunteers in UKB may lead to conservative estimates of penetrance. Third, though our sample size exceeded 72,000 exomes and enabled us to ascertain rare variants, many penetrance estimates for rare variants (e.g., singletons) are based on low numbers of carriers and may produce variable estimates. We included raw counts of carriers and non-carriers for each variant in the dataset so that estimates may be interpreted accordingly. Fourth, we mapped variants to diseases based on disease genes reported in ClinVar from pathogenic variant submissions. Inaccuracies in ClinVar submissions, and therefore our mapping from variant to disease, are possible, though we manually checked mappings extensively against the literature for accuracy.

In conclusion, we present a large-scale, systematic investigation of variant penetrance that leveraged thousands of exomes linked to the EHR. While we made available the penetrance measurements of all 37,772 variants for full transparency, we utilized a stringent set of 5,359 impactful variants that were ClinVar pathogenic or LoF in a non-recessive gene for many analyses. We demarcated differences in disease risk among variants of distinct ClinVar pathogenicity classes, review status, and molecular consequences. We also accessed a rich resource of clinical phenotypes to thoroughly explore penetrance: detailed physician notes, medications, biological measurements, and laboratory results. Critically, we investigated multiple dimensions of penetrance data spanning five ancestries and six decades of age thresholds. This study provides a blueprint for future studies to efficiently and accurately determine variant penetrance, the results of which will greatly improve our understanding of the genetic underpinnings of human disease.

## METHODS

### Study populations and sample filtering

A flowchart of the study design is provided (**Supplementary Figure 1**). We evaluated penetrance from individuals in two large-scale electronic health record (EHR)-linked population-based biobanks: The Bio*Me* Biobank (Bio*Me*) and UK Biobank (UKB). The study protocols were approved by the Institutional Review Board (IRB) of the Icahn School of Medicine at Mount Sinai. Use of data from UKB was completed and approved using the UK Biobank Resource under Application Number 16218. Informed consent was obtained for all study participants in both Bio*Me* and UKB through the approved IRB protocols. An overview of the demographics and clinical traits for both study populations is provided (**Table 1**). Bio*Me* is an EHR-linked biobank for ∼50,000 patients of African, Hispanic, European, and Other (Asian, Native American, and miscellaneous) self-reported ancestry who are recruited from the Mount Sinai Health System in Manhattan, NYC from 2007 onwards. All Bio*Me* participants consented to providing biological and DNA samples linked to de-identified EHRs. A subset of the individuals (n=31,250) was exome sequenced before undergoing quality control. A total of 229 samples with discordance between genetic sex and sex listed in the manifest, low coverage, contamination, low call rate, or duplications were excluded, leaving 30,813 samples. In addition, samples lacking complete demographic data (n=345), younger than 20 years of age (n=610), or without International Classification of Diseases-Clinical Modification 10 (ICD-10) diagnosis data (n=819) were removed to generate the final study set of 29,039 samples.

The UKB is a population-based longitudinal cohort of ∼500,000 individuals chiefly of British self-reported ancestry between 40-69 years of age who were enrolled at various sites across the United Kingdom between 2006-2010^21,67^. All individuals consented to providing medical history, demographic data, and DNA samples. A subset of 49,960 individuals had their exome sequenced and passed standard quality control (QC) filters, described extensively elsewhere^68^. We further excluded samples that lacked complete demographic information (n=2), or did not have ICD-10 diagnosis codes available (n=6,563), leaving a final study set of 43,395 samples for analysis.

### Whole-exome sequencing and quality control

In Bio*Me*, variant call files (VCFs) produced by Illumina v4 HiSeq 2500 contained 9,202,884 variants that were called in the samples. Goldilocks Filter (GF) was implemented on the VCFs. For single nucleotide polymorphisms (SNPs), cells with depth-normalized quality scores <3 or depth of coverage <7 were set to missing. For insertions and deletions (indels), cells with depth-normalized quality scores <5 or depth of coverage <10 were set to missing. Variant sites were then filtered, whereby sites of heterozygous variation failed the Allele Balance (AB) cutoff and were removed. SNP sites required ≥1 sample to carry an alternate AB ≥15% and indel sites required ≥1 sample to carry an alternate AB ≥20%. Together, these site filters removed 441,406 sites, leaving 8,761,478 variants after GF. Next, sites with missing genotypes for >2% of individuals in the dataset (267,955 sites) were removed. AB was calculated for biallelic SNPs and 320,877 sites with AB <0.3 or >0.8 were removed, leaving 8,172,646 sites. Lastly, the dataset was filtered to regions within the target regions of the exome capture platform (4,256,827 sites) and separated into 2 file sets for biallelic and multiallelic sites (3,948,623 and 308,204, respectively) due to differences in QC procedures.

In UKB, exome data from the first tranche of exome sequence data generated with the Functional Equivalence pipeline^68^ was used. Sequence data and QC for UKB are described elsewhere^21,67^.

### Variant curation and annotation

Using exome data from the 72,434 study samples, 37,772 variants associated with ClinVar^1^ diseases were ascertained using PLINK version 2.0^69^. To enrich for penetrant variants, we selected a strict subset of 5,359 clinically impactful (impactful) variants for most downstream analyses. These were defined by curating variant summary information in ClinVar VCF files released in December 2019, functional annotations from Variant Effect Predictor (VEP)^28^ version 99.2, and genic mode of inheritance from Online Mendelian Inheritance in Man (OMIM)^70^. An overview of variant selection is provided in **Supplementary Figure 2**. First, we included variants of pathogenic and/or likely pathogenic classification in ClinVar, and previously unreported variants with a damaging molecular consequence (splice acceptor/donor, stop gained/lost, frameshift, or start lost; collectively defined as LoF) annotated by VEP. LoF variants in a gene were mapped to disease based on prior pathogenic variant submissions in ClinVar linking genes to diseases (e.g., *BRCA1* LoF variants were mapped to breast cancer based on prior pathogenic variant submissions in ClinVar linking *BRCA1* to breast cancer). This filtered out benign variants or variants with uncertain or conflicting clinical significance in ClinVar and variants with synonymous molecular consequence. As missense variants have varying and uncertain degrees of pathogenicity, non-ClinVar missense variants were also excluded. We also noted the review status for each ClinVar variant, which summarizes the level of evidence supporting a variant’s claim of clinical significance ranging from the lowest level of no assertion criteria (review status=0), to single submitter or multiple submitters with conflicting interpretation (review status=1), to multiple submitters with no conflict of interpretation (review status=2), to reviewed by an expert panel such as ClinGen^71^ (review status=3), to the highest level of practice clinical guidelines (review status=4). Second, we excluded variants in genes with exclusively recessive mode of inheritance reported in OMIM. The gene for each variant was retrieved from NCBI reference sequences (RefSeq^72^) and corroborated with genomic coordinates in OMIM. The mode of inheritance was mined for all phenotypes reported for each gene in OMIM and genes were then summarized as Dominant (only dominantly inherited phenotypes), Both (both dominantly and recessively inherited phenotypes), or Recessive (only recessively inherited phenotypes).

### Phenotyping of carriers

In both UKB and Bio*Me*, we obtained case status using ICD-10 diagnosis codes. ICD-10 codes are commonly used in genetic population studies to define cases of a disease^61,62^ and they map directly to ClinVar diseases in the Systematized Nomenclature of Medicine Clinical Terms (SNOMED CT)^27^. All of the 29,039 Bio*Me* samples and 43,395 UKB samples in the final dataset had ICD-10 diagnosis codes available. Cases were identified by the presence of a corresponding ICD-10 code while controls were identified by the absence of all corresponding ICD-10 codes. We thereby defined case-control status for over 400 SNOMED CT diseases of non-recessive inheritance in ClinVar (disease inheritance was retrieved from NCBI’s MedGen database at https://www.ncbi.nlm.nih.gov/medgen/); 197 of the diseases were present with at least one case in the final dataset. A complete list of the cases, controls, and ICD-10 diagnosis codes for each disease is provided in **Supplementary Table 1**.

Validation of the ICD-10-based phenotyping method was performed in a set of three analyses in Bio*Me*. First, penetrance estimates using the ICD-10 phenotypes were compared against results using clinical algorithms from the literature for nine diseases^29–37:^ age-related macular degeneration (AMD), arrhythmogenic right ventricular cardiomyopathy (ARVC), Brugada syndrome (BrS), familial hypercholesterolemia (FaH), familial breast cancer (FBC), hepatocellular carcinoma (HCC), idiopathic pulmonary arterial hypertension (IPAH), prostate cancer (PCa), and type 2 diabetes (T2D) (**Supplementary Table 3**, **Supplementary Table 4**). Second, we verified ICD-10 phenotypes by manually reviewing physician notes in the problems list (PL) for six diseases: Alzheimer’s disease (AD), AMD, FaH, FBC, ischemic stroke, and T2D (**Supplementary Table 5**). For each disease, we extracted physician notes for a random sample of 50 ICD-10-based cases and 50 ICD-10-based controls. The presence of symptoms or findings indicative of a diagnosis corroborated ICD-10-based cases, while the absence of symptoms or findings corroborated ICD-10-based controls. Third and lastly, we examined the association of biomarker levels with penetrance estimates for variants in genes with clear biological roles in three diseases: *LDLR*^12^ in FaH (measured by LDL-C and total cholesterol), *HNF1A*^38^ in MODY (measured by glucose and hemoglobin A1c), and *UCP3*^39,40^ in obesity (measured by BMI and weight). We extracted median biological measurements for carriers and medication usage relevant to FaH (statins) and T2D (insulin, insulin analogs, pramlintide, glucagon-like peptide 1 agonists, metformin, sulfonylurea, dipeptidyl peptidase 4 inhibitors, glitazone, sodium-glucose transport protein 2 inhibitors, or alpha-glucosidase inhibitors) to control for the effect of medications on biological measurements in association analyses.

## Statistical analysis

All statistical tests and plots were made using R statistical software version 3.5.3^75^. Differences in categorical variables were assessed with a Fischer’s exact test, while differences in continuous variables were tested with a two-sided t-test. Significance level was set at *P*<0.05 for comparisons between two groups. Risk difference (RD) between the prevalence of disease in carriers and non-carriers was computed, and significance was evaluated with Fisher’s exact test. A strict Bonferroni correction was applied to *P*-values in analyses involving multiple comparisons, including comparisons of mean penetrance for five ClinVar pathogenicity classes, mean penetrance for four ClinVar review status levels, and mean penetrance for eight molecular consequences. Phenotype validation using PL physician notes was performed by computing the ratio of PL-based cases to ICD-10-based cases and PL-based controls to ICD-10-based controls to determine case and control concordance, respectively. Association analyses of variant penetrance with biomarker levels were performed using a multivariable linear regression, adjusted for clinical covariates of age, sex, BMI (except in the analysis of obesity), 10 genetic principal components (PCs), and medication usage where relevant.

## Data Availability

The UKB data may be browsed at http://biobank.ndph.ox.ac.uk/showcase/ and access to data can be requested at https://www.ukbiobank.ac.uk/register-apply/. More information about BioMe can be found at https://icahn.mssm.edu/research/ipm/programs/biome-biobank/researcher-faqs. The complete penetrance dataset used for all analyses is provided (Supplementary Tables 9 and 10) with no restrictions on the data released.

## Data availability

The UKB data may be browsed at http://biobank.ndph.ox.ac.uk/showcase/ and access to data can be requested at https://www.ukbiobank.ac.uk/register-apply/. More information about Bio*Me* can be found at https://icahn.mssm.edu/research/ipm/programs/biome-biobank/researcher-faqs. The complete penetrance dataset and gene disease map used for all analyses (**Supplementary Tables 1, 9** and **10**) will be made available upon peer review with no restrictions on the data released.

## Author Contributions

IF and RD designed the study. IF, RL, JC, and RD obtained the data. IF, KC, HV, SB, DJ, and GR analyzed and/or interpreted the data. IF and RD drafted and revised the manuscript. All authors read and provided critical comments on the manuscript.

## Acknowledgements

The Bio*Me* healthcare delivery cohort at Mount Sinai was founded and maintained with a generous gift from the Andrea and Charles Bronfman Philanthropies. We thank the individuals who were involved in the quality control and/or file handling for the exome sequencing and genome-wide genotyping data, including Aayushee Jain, Lisheng Zhou, Michael Preuss, Quingbin Song, Stephane Wenric, and Steve Ellis. We also thank the thesis advisory committee of IF, including Bruce D. Gelb, Sander Houten, Paz Polak, and Stuart Scott, for their critical feedback and expertise. Research reported in this paper was supported by the Office of Research Infrastructure of the National Institutes of Health under award numbers S10OD018522 and S10OD026880. The content is solely the responsibility of the authors and does not necessarily represent the official views of the National Institutes of Health.

