## Supplementary Figure for "Ancestrally and Temporally Diverse Analysis of Penetrance of Clinical Variants in 72,434 Individuals"

### Supplemental Material

### FIGURES

**Supplementary Figure 1**. Graphical abstract of study design and workflow.

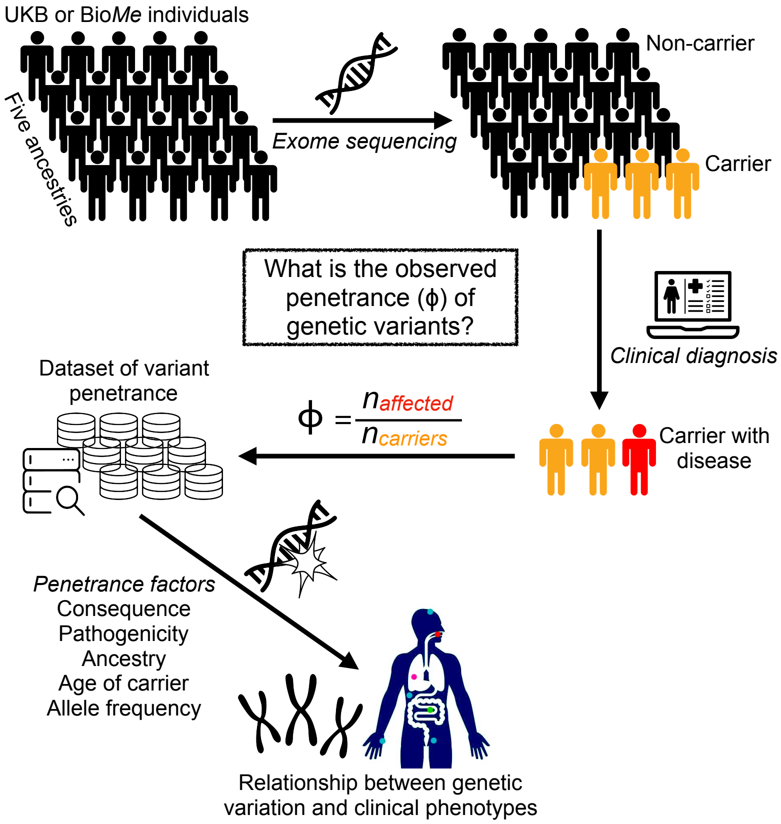


Graphical abstract of study design and workflow. A total of 72,434 individuals from the Bio*Me* Biobank (Bio*Me*) and UK Biobank (UKB) with exome sequence data were used to analyze penetrance of 37,772 clinical variants. Genetic information was linked to clinical phenotypes in the electronic health record (EHR), including diagnosis codes, metabolite levels, biological measurements, and physician notes, and was used to compute penetrance. The results were used to examine how penetrance varies by ClinVar pathogenicity class, molecular consequence, and carrier age and ancestry.

**Supplementary Figure 2**. Variant selection for analysis of penetrance.

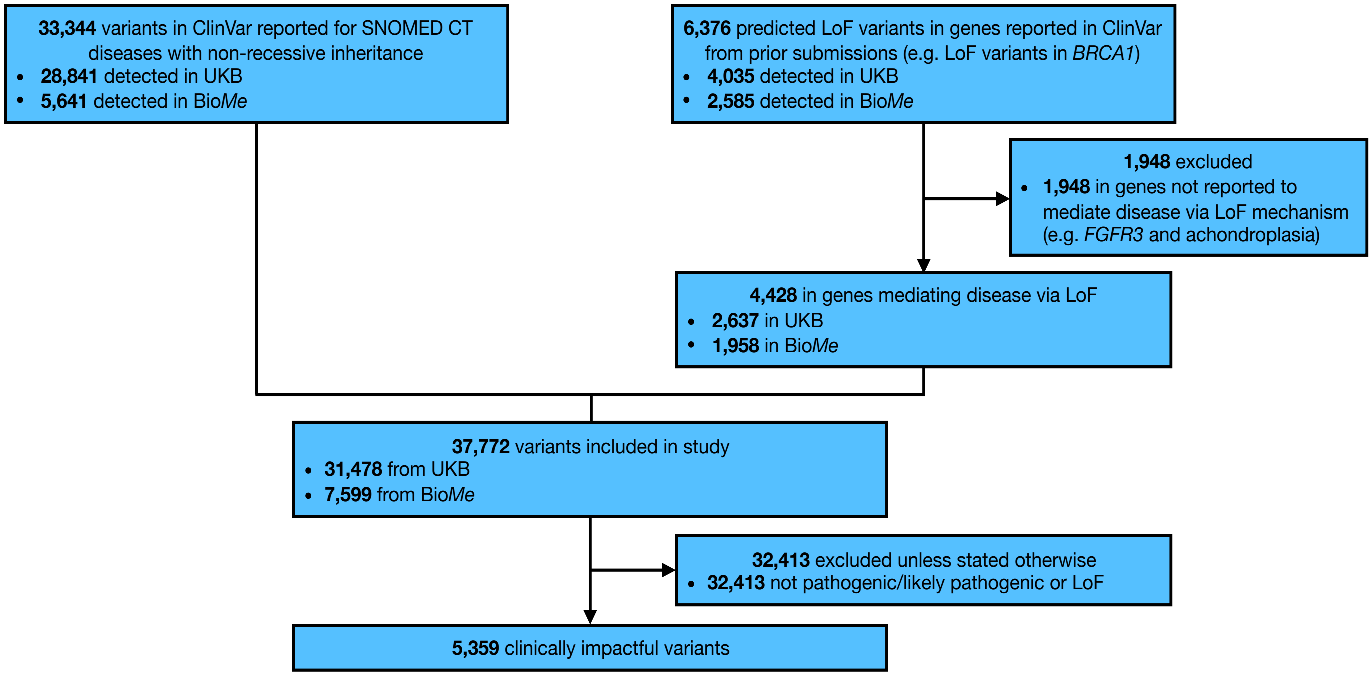


Selection criteria of genetic variants included for analysis of penetrance in the Bio*Me* Biobank (Bio*Me*) and UK Biobank (UKB). Loss-of-function (LoF) variants in a gene were mapped to disease based on prior variant submissions in ClinVar linking the gene to disease (e.g., LoF variants in *BRCA1* were mapped to breast cancer based on prior variant submissions in ClinVar linking *BRCA1* variants to breast cancer). LoF variants in genes that do not mediate disease via LoF mechanism were excluded (e.g., *FGFR3* in achondroplasia). Clinically impactful variants are reported as pathogenic/likely pathogenic in ClinVar or are of LoF consequence in a gene mediating disease via LoF mechanism, and in a gene with non-recessive inheritance.

**Supplementary Figure 3**. Sensitivity analysis of penetrance estimates by sample size.

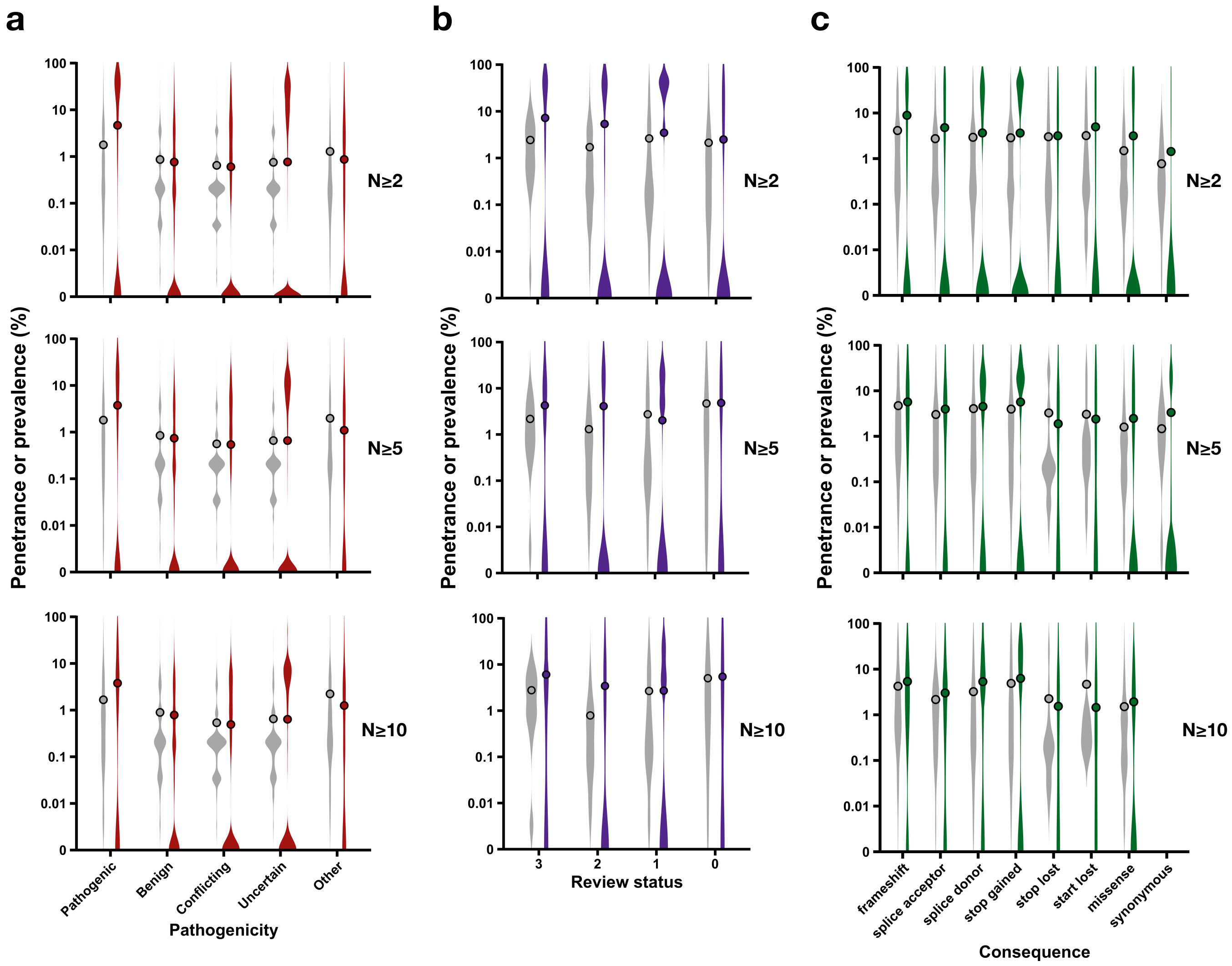


Sensitivity analysis of penetrance estimates by sample size. For increasing thresholds of sample size (N≥2 carriers [excludes singletons], N≥5 carriers, and N≥10 carriers), penetrance distributions are shown as violin plots in red, purple, and green color on a base-10 logarithmic scale with the mean penetrance overlaid as points, alongside disease prevalence in non-carriers shown as violin plots in grey color with the mean disease prevalence superimposed as points. Pathogenic/likely pathogenic variants are grouped as pathogenic and benign/likely benign variants are grouped as benign. **a**,

Penetrance is stratified by classification of variant pathogenicity in ClinVar. Pathogenic variants on average have the highest penetrance for all sample size thresholds (4.7% vs. 0.76% next highest mean for uncertain class [N≥2], 3.7% vs. 1.1 % next highest mean for other class [N≥5], and 3.8% vs. 1.3% next highest mean for other class [N≥10]; two-tailed t-test $P$=5x$10^{-134}$, $P$=1x$10^{-18}$, and $P$=1x$10^{-15}$, respectively). Other pathogenicity, variants with a ClinVar pathogenicity classification other than pathogenic, benign, conflicting, or uncertain. **b**, Penetrance is stratified by variant review status in ClinVar as reviewed by experts (review status=3), multiple submitters (review status=2), single submitter (review status=1), or no assertion criteria (review status=0). Variants reviewed by experts (review status=3) have the highest penetrance on average for all sample size thresholds (7.2% vs. 5.4% next highest review status=2 group [N≥2], 4.3% vs. 4.1% next highest review status=2 group [N≥5], and 6.0% vs. 3.4% next highest review status=2 group [N≥10]; $P$=2x$10^{-1}$, $P$=9x$10^{-1}$, and $P$=3x$10^{-1}$, respectively). **c**, Penetrance is stratified by molecular consequence using Variant Effect Predictor (VEP). Frameshift variants on average have the highest penetrance for all sample size thresholds (8.9% vs. 3.2% for missense [N≥2], 5.7% vs. 2.5% for missense [N≥5], and 5.4 vs. 1.9% for missense [N≥10]; $P$=6x$10^{-10}$, $P$=5x$10^{-3}$, and $P$=1x$10^{-2}$, respectively).

**Supplementary Figure 4**. Penetrance of 13,298 singletons stratified by ClinVar pathogenicity, ClinVar review status, and molecular consequence.

**Penetrance dataset** (n=37,772 variants [197 diseases])

**Variants in non-recessive genes** (n=34,301 [181])

**Singletons** (n=13,298 [175])

**In ClinVar** (n=10,487 [157])

**Impactful** (n=3,507 [154])

**Stratify by ClinVar pathogenicity**

| Pathogenicity | Singletons | Penetrant | Proportion penetrant (%) |
|---|---|---|---|
| Pathogenic | 906 | 120 | 13 |
| Benign | 2,374 | 54 | 2.3 |
| Conflicting | 1,113 | 29 | 2.6 |
| Uncertain | 5,992 | 116 | 1.9 |
| Other | 102 | 1 | 0.98 |

**Pathogenic** (n=906 [112])

**Stratify by ClinVar review status**

| Review status | Singletons | Penetrant | Proportion penetrant (%) |
|---|---|---|---|
| 3 | 115 | 47 | 41 |
| 2 | 362 | 42 | 12 |
| 1 | 319 | 24 | 7.5 |
| 0 | 109 | 7 | 6.4 |

**Stratify by molecular consequence**

| Consequence | Singletons | Penetrant | Proportion penetrant (%) |
|---|---|---|---|
| frameshift | 759 | 119 | 16 |
| splice acceptor | 338 | 22 | 6.5 |
| splice donor | 469 | 24 | 5.1 |
| stop gained | 1,541 | 107 | 6.9 |
| stop lost | 19 | 0 | 0 |
| start lost | 68 | 0 | 0 |
| missense | 293 | 23 | 7.8 |
| synonymous | 7 | 0 | 0 |

Penetrance of 13,298 singletons stratified by ClinVar pathogenicity, ClinVar review status, and molecular consequence. Review status, level of evidence for a variant's assertion of pathogenicity in ClinVar ranging from 0 (no assertion criteria) to 3 (reviewed by experts). Clinically impactful variants are reported as pathogenic/likely pathogenic in ClinVar or are of loss-of-function consequence in a gene mediating disease via LoF mechanism, and in a gene with non-recessive inheritance.

**Supplementary Figure 5**. Association between age of disease onset and age-dependent change in penetrance for 157 diseases in the Bio*Me* Biobank and UK Biobank.

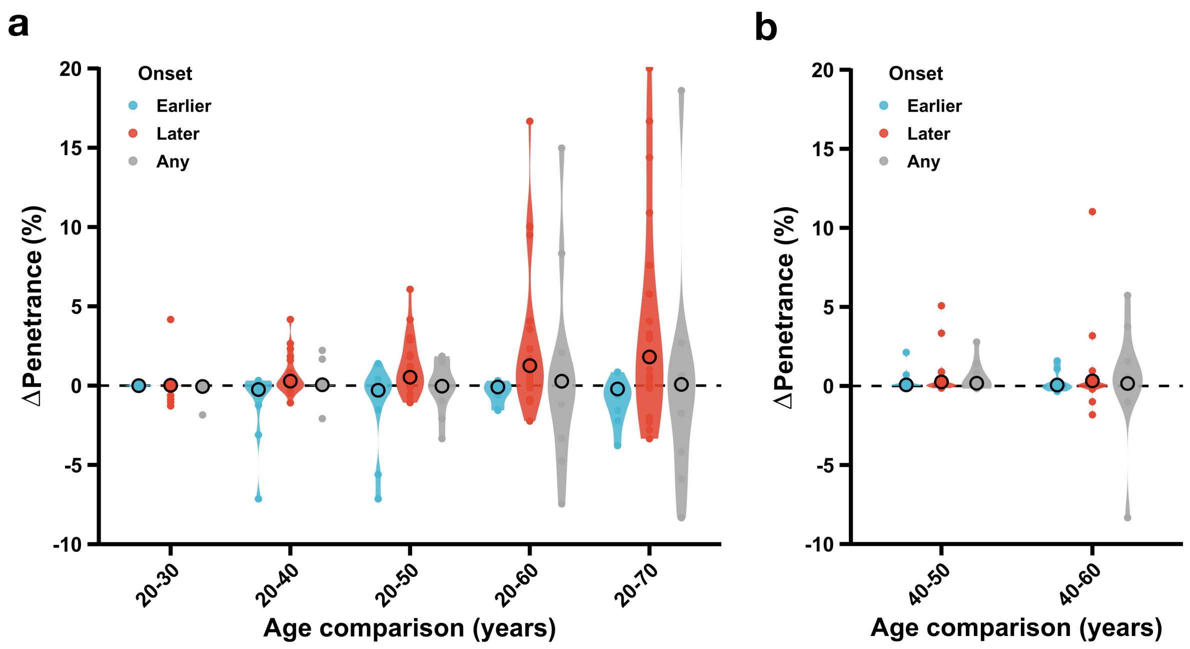


Association between age of disease onset and age-dependent change in penetrance for 157 diseases in the Bio*Me* Biobank and UK Biobank. Diseases correspond to the 5,359 clinically impactful variants and are grouped according to their age of onset: Earlier, Later, or Any. Change in penetrance is displayed as a violin plot for each age of onset group when comparing two carrier age thresholds with the mean change in penetrance superimposed as a point. ΔPenetrance (%), change in variant penetrance represented as a percent (+ values indicate greater penetrance estimate with the older age threshold and - values indicate greater penetrance estimate with the younger age threshold); age comparison, two carrier age thresholds for which penetrance is compared (e.g. 20-70 compares penetrance with carriers ≥20 years old and penetrance with carriers ≥70 years old); onset, disease groups according to age of onset. **a**, The Bio*Me* Biobank has carriers of age 20-90 years old and ΔPenetrance is assessed over five carrier age thresholds: Later diseases have greater ΔPenetrance on average than Earlier diseases for 20-40 (two-tailed t-test *P*=0.006), 20-50 (*P*=0.002), 20-60 (*P*=0.02), and 20-70 (*P*=0.02) age comparisons. **b**, UK Biobank has carriers of age 40-69 years old and ΔPenetrance is evaluated over two carrier age thresholds: there is no difference in ΔPenetrance between Later and Earlier diseases for either 40-50 (*P*=0.2) or 40-60 (*P*=0.2) age comparisons.

### TABLES

**Supplementary Table 1**. Summary of cases/controls, ICD-10 diagnosis codes, and genes for 197 diseases analyzed for penetrance of genetic variants.

**Supplementary Table 2.** Summary of 37,772 clinical variants assessed for penetrance.

**Supplementary Table 3.** Validation of ICD-10-based phenotypes with clinical algorithms in the assessment of penetrance for nine diseases in the Bio*Me* Biobank.

**Supplementary Table 4**. Tabulated list of 208 penetrance measurements computed with ICD-10-based phenotypes and clinical algorithms for ClinVar pathogenic variants in the Bio*Me* Biobank.

**Supplementary Table 5.** Validation of ICD-10 phenotypes with manual curation of physician notes in the problem list (PL) for six diseases in the Bio*Me* Biobank.

**Supplementary Table 6.** Summary of risk difference for 5,359 clinically impactful variants across 197 diseases.

**Supplementary Table 7.** Age of onset for 197 diseases included in study.

**Supplementary Table 8.** Examples of 20 clinically impactful variants associated with elevated risk of seven diseases.

**Supplementary Table 9.** Penetrance dataset for 12,844 variants analyzed in the Bio*Me* Biobank.

**Supplementary Table 10.** Penetrance dataset for 60,407 variants analyzed in the UK Biobank.
